# Development and Prospective Validation of Predictive Model for Early Hemodynamic Deterioration in Critical Care: A Multicenter Study

**DOI:** 10.64898/2026.06.05.26353765

**Authors:** Aditya Nagori, Pradeep Singh, Sameena Firdos, Akshaya Devadiga, Vanshika Vats, Arushi Gupta, Harsh Bandhey, Prakriti Ailavadi, Raghav Awasthi, N Narotam, Aditya Mishra, Rakesh Lodha, Tavpritesh Sethi

**Author notes:** Corresponding author. Prof. Tavpritesh Sethi. E-mail(s). These authors contributed equally to this work (Aditya Nagori, Pradeep Singh).

## Abstract

High-frequency physiological monitoring in ICUs can identify impending deterioration hours before clinical recognition yet extracting reliable early-warning signals from noisy vital-sign streams remains challenging. We present SIgnose, an interpretable prediction framework for early detection of abnormal shock index (SI), built from routinely monitored vital signs using physiologic variability and nonlinear time-series features. SIgnose was developed on the eICU Collaborative Research Database and externally validated on the MIMIC-III adult database and a pediatric SafeICU cohort (AIIMS New Delhi), with additional prospective validation in the pediatric ICU.

We benchmarked three representation strategies: (i) engineered physiologic variability and nonlinear time-series features, (ii) deep learning, and (iii) Llama-3.1-8B embeddings with low-rank adaptation. Physiologic variability features consistently demonstrated superior cross-cohort generalization. The final model used 3,970 features from five vital signs to predict abnormal SI up to 8 hours ahead, achieving AUROC 0.861 (95% CI 0.859-0.863) and AUPRC 0.927 (95% CI 0.925-0.929) on eICU. External validation yielded AUROC 0.870 (95% CI 0.863-0.876) and AUPRC 0.935 (95% CI 0.930-0.940) on MIMIC-III, and AUROC 0.875 (95% CI 0.863-0.888) and AUPRC 0.915 (95% CI 0.898-0.930) on SafeICU; prospective pediatric validation (n = 88) achieved AUROC 0.885 (95% CI 0.868-0.902) and AUPRC 0.911 (95% CI 0.882-0.936).

SHAP interpretability analysis identified heart rate variability, respiratory trend dynamics, and multi-scale blood pressure variability as key early-warning signatures. These findings establish SIgnose as a reproducible, low-compute, early-warning framework and demonstrate that physiologic variability features provide robust, generalizable representations for early deterioration detection across adult and pediatric critical care.

## INTRODUCTION

Delays in recognizing physiological deterioration in intensive care units (ICUs) can rapidly precipitate irreversible organ dysfunction and excess mortality (1). Shock in ICUs has a substantial global burden with mortality as high as 34% in low- and middle-income settings, with septic shock mortality approaching 38% even in high-income regions (2–4). Because earlier intervention can avert decompensation and downstream multi-organ failure, reliable early identification of hemodynamic instability remains a central objective of critical care (1,5–8). A key barrier to developing a scalable and generalizable early-warning system for such decompensation is the dependence of many predictive models on laboratory values, clinician charting, or sparse electronic health record (EHR) features. These data streams are often heterogeneous across sites, intermittent, and delayed in relation to disease progression. In contrast, bedside monitors provide a ubiquitous, high-frequency stream of vital signs (e.g., heart rate, blood pressure, respiratory rate, oxygen saturation) that is available across health systems and care environments, including resource-limited settings (9,10). These signals contain rich information about autonomic regulation, compensatory dynamics, and multiscale variability that can precede overt decompensation. However, learning early-warning models from continuous monitoring remains challenging due to noise, missingness (11), variable temporal resolution, and dataset shift across hospitals (12), age groups, and monitoring practices (13).

Shock Index (SI), the ratio of heart rate to systolic blood pressure, is a simple, unitless bedside marker that requires no clinician judgment and can be computed continuously from standard ICU monitors (14–15). Originally proposed as a measure of hemodynamic stability, SI has since demonstrated broad prognostic and risk-stratification utility across diverse clinical syndromes and settings, including sepsis, pneumonia, acute myocardial infarction, stroke, advanced cancer, pulmonary embolism, and trauma (2,14–15). This breadth makes SI especially attractive as an early-warning target: it reflects a physiologic “final common pathway” of stress and circulatory compromise that can emerge across multiple disease etiologies, allowing a single endpoint to support general-purpose deterioration surveillance in critical care. Moreover, abnormal SI exposure is itself associated with clinically meaningful outcomes in critically ill populations, including adverse outcomes and mortality (16). These features make SI not only clinically interpretable and operationally simple, but also potentially more transportable across institutions than endpoints requiring lab thresholds or site-specific treatment patterns.

A growing body of work applies Machine Learning (ML) and Deep Learning (DL) to ICU prediction tasks (17,18). In parallel, foundation models such as Llama-3.1-8B have enabled emerging strategies for representation learning by converting structured clinical time series into textual prompts and embeddings (19–20). The generalizability of predictions remains a major challenge when transported across hospitals, age groups, and health systems due to dataset shift, heterogeneous temporal resolutions, differences in onset definitions, variable measurement practices, and missingness patterns. This motivates a direct comparison of foundation-model embeddings against representations grounded in physiological variability and nonlinear time-series dynamics. We hypothesized that physiological features from monitor-only vital sign streams can achieve robust multicenter generalization for the early prediction of abnormal SI, provided that appropriate temporal representations are learned (9–10). Importantly, such a low-compute, monitor-only approach facilitates real-time bedside deployment and enables use in settings where access to GPUs, cloud-based inference, or proprietary foundation models is limited, thereby supporting more equitable dissemination of early-warning systems across diverse critical care environments.

To test the hypothesis in a comparative setting, we developed and benchmarked a physiological feature-based model, comparing it with embeddings from Meta’s Llama-3.1-8B, a strong open weight baseline foundation model. Our model was developed using the eICU (a multicenter ICU database) (12) and externally validated on the Medical Information Mart for Intensive Care-III (MIMIC-III) adult dataset (11) and a novel SafeICU pediatric dataset from All India Institute of Medical Science (AIIMS), New Delhi (21). We additionally performed prospective validation in the pediatric ICU setting. Figure 1 summarizes the study design and evaluation strategy.

**Figure 1:**
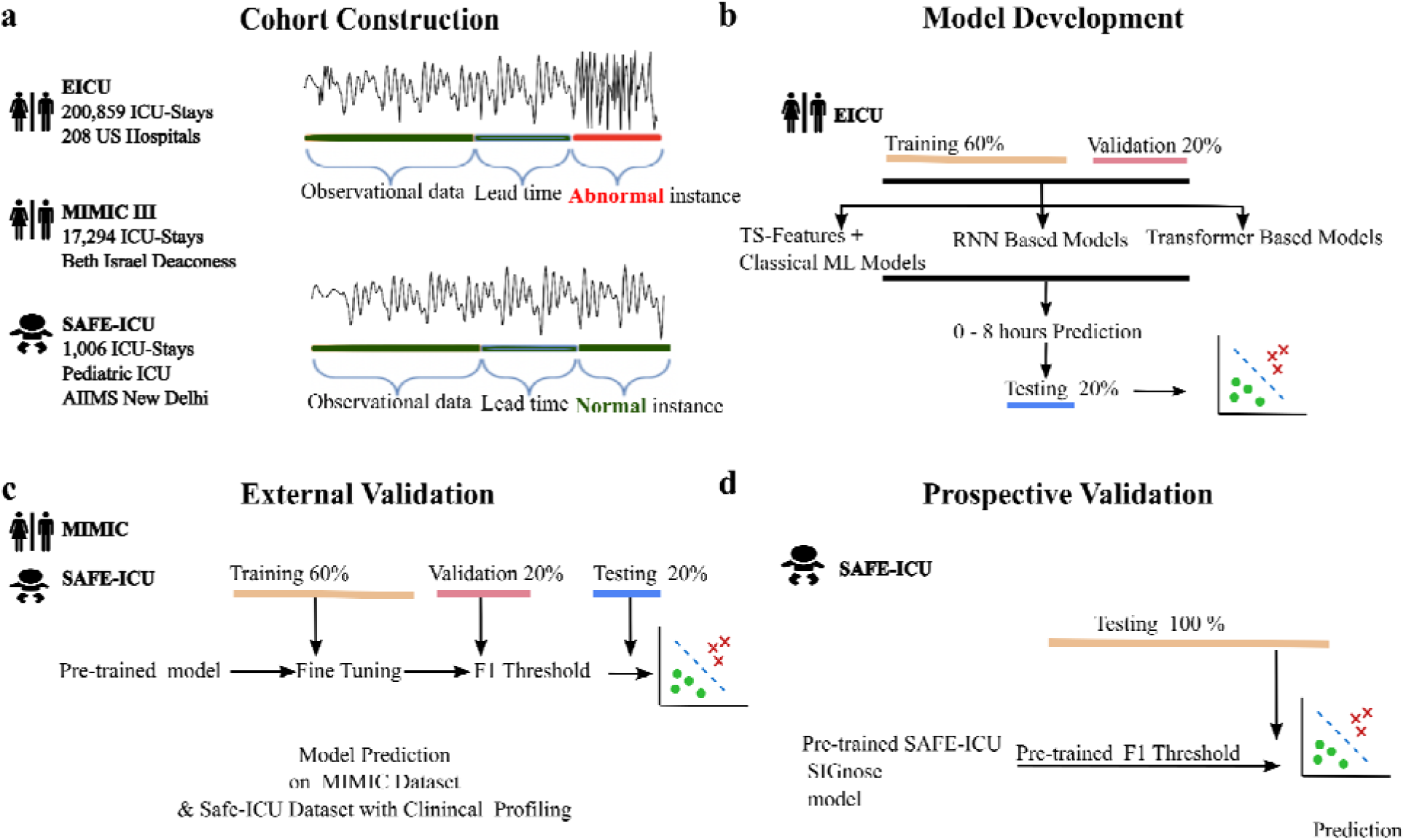
Graphical Abstract: Summary of the pipeline for generalizing the prediction of hemodynamic deterioration. 1a) shows cohort construction, where a physiological time series of *heart rate, systolic arterial blood pressure, diastolic arterial blood pressure, respiratory rate,* and *oxygen saturation* was used. 1b) shows the pipeline for constructing models to predict hemodynamic deterioration 0-8 hours in advance and for evaluation. 1c) shows the generalization of our best model, *SIgnose* learned models on MIMIC-III data and Indian settings’ SafeICU pediatric data. 1d) shows the prospective validation on the prospective SafeICU pediatric dataset.

Our contributions are threefold: (i) a head-to-head evaluation of ML, DL, and Llama-3.1-8B-based embedding strategies for monitor-only early prediction of abnormal SI from high-frequency vital-sign streams; (ii) evidence that physiologic variability feature-based time-series models provide superior transportability relative to embedding-based LLM textualization across heterogeneous ICU cohorts; and (iii) the development and external and prospective validation of a clinically practical early-warning system, SIgnose, designed for bedside integration to reduce delays in recognition and treatment of hemodynamic deterioration.

## METHODS

### Dataset description & pre-processing

We leveraged two publicly available datasets from the USA and a SafeICU pediatric in-house dataset with 0.23 million patient hours from a tertiary care setting in India. The multicenter eICU data, comprising 200,859 unit admissions, of which 192,751 contained physiological time series data, collected from 208 hospitals in 2014 to 2015 (12), were used to construct the cohort. For external validation, we used (i) a SafeICU pediatric dataset warehoused from February 2016 to January 2020, containing 1006 ICU stays from 782 patients. (ii) MIMIC-III v1.4 data collected between 2001 and 2012. The MIMIC-III physiological time series includes records for the adult population, including 17,294 ICU stays involving 10,269 patients. In addition to external validation, data collected from February 2020 to August 2020 in the pediatric ICU were used for prospective validation and generalization of models developed on eICU data.

### Cohort Construction Development cohort

Electronic ICU (eICU) data were used to construct the model development cohort, which included data categorized as medical (MICU), surgical (SICU), medical-surgical ICU, coronary care (CCU), cardiac ICU, cardiac surgery *(CSICU)*, *cardiothoracic (CTICU)*, and *neuro-ICU*. Our inclusion criteria for the development cohort included all patients with continuous vital sign monitoring for at least 7.5 hours during their ICU stay. The exclusion criteria were the following: (i) patients with non-availability of invasive arterial blood pressure monitoring in the observation window, (ii) patients with non-availability of patient demographics (age, gender, or age greater than 89 years), (iii) patients with abnormal SI within the first 7 hours of admission to avoid biasing the model with patients already in shock, and finally (iv) patients with missing data in >10% of the observation period.

### External validation cohorts

MIMIC-III and SafeICU data were used as external validation cohorts, with inclusion and exclusion criteria like those of the development cohort. Since SafeICU is a pediatric data resource, different SI thresholds were used for various age groups (SI >2.3 (ages <= 3 months), >1.7 (4-6 months), >1.5 (7-12 months), >1.2 (13-36 months), >1.15 (37-72 months), >0.95 (73-144 months), >0.77 (>144 months))(22).

Prospective validation cohort. Prospective validation cohorts were constructed using prospectively collected data from the pediatric ICU facility at AIIMS, New Delhi (n = 88; February–August 2020), using the same inclusion and exclusion criteria.

### Pre-processing and Scoring

Heart rate, systolic arterial blood pressure, diastolic arterial blood pressure, respiratory rate, and oxygen saturation were used as physiological vitals in the model. Minimal preprocessing of the vital signs data was performed. Critical patient data for eligible patients in the development cohort were imputed using the Kalman filter. The SI score was calculated for each timestamp in the preprocessed time series. To label the physiological time series for SI clinical deterioration, we divided each physiological time series into 30-minute epochs. The onset time of abnormal SI (t _(ASI)_) is defined as the starting time of an epoch in which the median SI value was greater than 0.7 (8,21). The training window was 7 hours, with the index time defined as the onset time plus a lead time of up to 8 hours (Supplemental Material 1). Exploratory data analyses were conducted in R version 3.6 to characterize the data and assess associations between the first three days of SI status, length of stay (LoS), and mortality incidence.

### Engineering Explainable Feature and Feature Selection

A set of 3970 time-series features (TS-features), including linear and nonlinear physiological features such as statistics, periodicity, wavelet, entropy, and other complex features, was extracted using the *tsfresh* (23) library. TS-features are explainable and capture physiological-domain understanding, unlike black-box features from DL. Variable selection was performed using a random forest-based Boruta algorithm (24), implemented in R.

### Model Development, Evaluation, and Interpretability

The development cohort consisted of patients who had abnormal SI within the next 8 hours of the index time. The outcome variable- abnormal SI within the next 8 hours was defined by SI scores. Since generalizability requires the models to perform across different sites, the training and testing sets were designed to contain non-overlapping clinical sites. Five-fold cross-validation sets were constructed, and hyperparameter tuning was performed using grid search to optimize performance on the validation set. F1 thresholds were used as probability cutoffs derived from the validation set. Results were reported as the median and standard deviation on fivefold test sets. The area under the receiver operating characteristic curve (AUROC), the area under the precision recall curve (AUPRC), and Recall & Precision were evaluated on the test set. Since only a small subset of patients undergoes invasive arterial blood pressure monitoring, two variants of the model (with and without invasive monitoring) were constructed and compared on the same sample size. The pre-trained model’s output threshold was tuned on the validation set to improve generalization to unseen data. Model performance for different types, i.e., medical, coronary care, surgical, cardiac surgery, and cardiothoracic ICUs, was evaluated. A comparison of model performance indicators for different lead times, age groups, and time since admission was also done.

Further, we compared standard ML models with feature engineering with Long Short-Term Memory (LSTM) (25) models without feature engineering. For the first approach, vital TS-features were computed, z-score normalized, and used to train gradient boosting machine (GBM) (26), random forest (27), and multilayer perceptron models using the sklearn (28) library in Python version 3.6.3. The catalogue of TS-features includes statistical, frequency domain, and nonlinear features (Supplemental Material 2). In the second approach, Bidirectional LSTM (Bi-LSTM) (29) and Convolutional Neural Network LSTM (CNN-LSTM) models were fitted to the raw physiological time series in Python 3.6 using Keras (version 2.3.1) and TensorFlow (version 2.1.0), and a softmax activation was used to generate probability scores. In the third approach, we used the Llama-3.1-8B model (30) with 4-bit quantization. We accessed it via the “unsloth” Python library, using a low-rank adapter (LoRA) (31) (Supplemental Material 13) with structured physiological time-series data to evaluate LLMs for shock prediction. Every patient record contained demographic information (age and gender) and physiological variables recorded every five minutes. These physiological variables included oxygen saturation, heart rate, respiratory rate, and arterial systolic and diastolic blood pressure. We prepared the data and converted it into structured text prompts that define patient physiological time series. The prompts followed a standardized instruction-based format and were fed to Llama-3.1-8B to generate contextual embeddings for each patient’s physiological state.

The prompt template was as follows: “Instruction: Analyze the following medical data and predict the outcome. Input: Age: {age}, Gender: {gender}, this data includes observations at 5-minute intervals containing—Oxygen saturation (%): {os2}, Heart rate (bpm): {hrate}, Respiratory rate (brpm): {resp_rate}, Arterial systolic blood pressure (mmHg): {ss}, Arterial diastolic blood pressure (mmHg): {sd}” These embeddings were used as features in multiple ML models: Support Vector Machine (SVM), XGBoost, LightGBM, and AdaBoost for the downstream classification task of predicting clinical shock. This setup allowed us to compare the embedding-based approach with traditional time-series modeling techniques.

Age and gender were included as covariates in all the approaches. The winning strategy and the fine-tuned models were tested for generalization and prospective validation. The Enhancing the Quality and Transparency of Health Research (EQUATOR) Network’s Transparent Reporting of a multivariable prediction model for Individual Prognosis or Diagnosis (TRIPOD) (32) guidelines were followed for developing the ML models and reporting results.

#### SHAP value computation for model interpretability

SHAP (SHapley Additive exPlanations) is a method that assigns each feature an importance value based on game-theoretic principles (33). SHAP values represent the influence of features on the model’s prediction. The SHAP value distributions for each feature across all test sets were computed. All test sets were combined to plot the model’s explanation of the top 15 most important features and their relationships with SHAP values.

### Model Generalization & Prospective Validation

SIgnose model performance was validated on publicly available retrospective MIMIC-III and in-house Pediatric data for external validation. Prospective validation was performed on in-house pediatric data. Both datasets were pre-processed to mitigate dataset-specific variation, including time-resolution adjustments and HL7 data cleaning. The high-resolution MIMIC-III and SafeICU data were summarized to a 5-minute resolution to align with the requirements of our model, which was trained on eICU data. For each dataset, three modeling scenarios were compared for validation: (i) with fine-tuning of parameters, (ii) without fine-tuning, and (iii) a siloed model, where the model was trained and evaluated using data from the same clinical site only, without incorporating data from other sites. Finally, prospective validation of the model was performed on the SafeICU data collected from February 2020 to August 2020.

## RESULTS

### Data characteristics

The development cohort comprised patients from the eICU database across various ICU types, including MICU, SICU, CCU, and others, with at least 7.5 hours of continuous vitals monitoring. External validation was performed on both adult (MIMIC-III) and pediatric (SafeICU) datasets, using consistent inclusion and exclusion criteria. Additionally, a prospective validation cohort was constructed using pediatric ICU data from AIIMS, New Delhi, ensuring applicability across age groups and real-world clinical settings.

The constructed cohort from the eICU database consisted of 16,246 ICU stays, of which 12,109 had at least one episode of abnormal SI (Supplemental Material 3a). The higher prevalence of abnormal SI in this cohort reflects the development cohort criteria, which include arterial blood pressure for assigning ground-truth labels. The median LoS for patients was 3.1 days, whereas in the abnormal SI, it was significantly higher at 4.28 (6.79) days (Table 1).

**Table 1:**
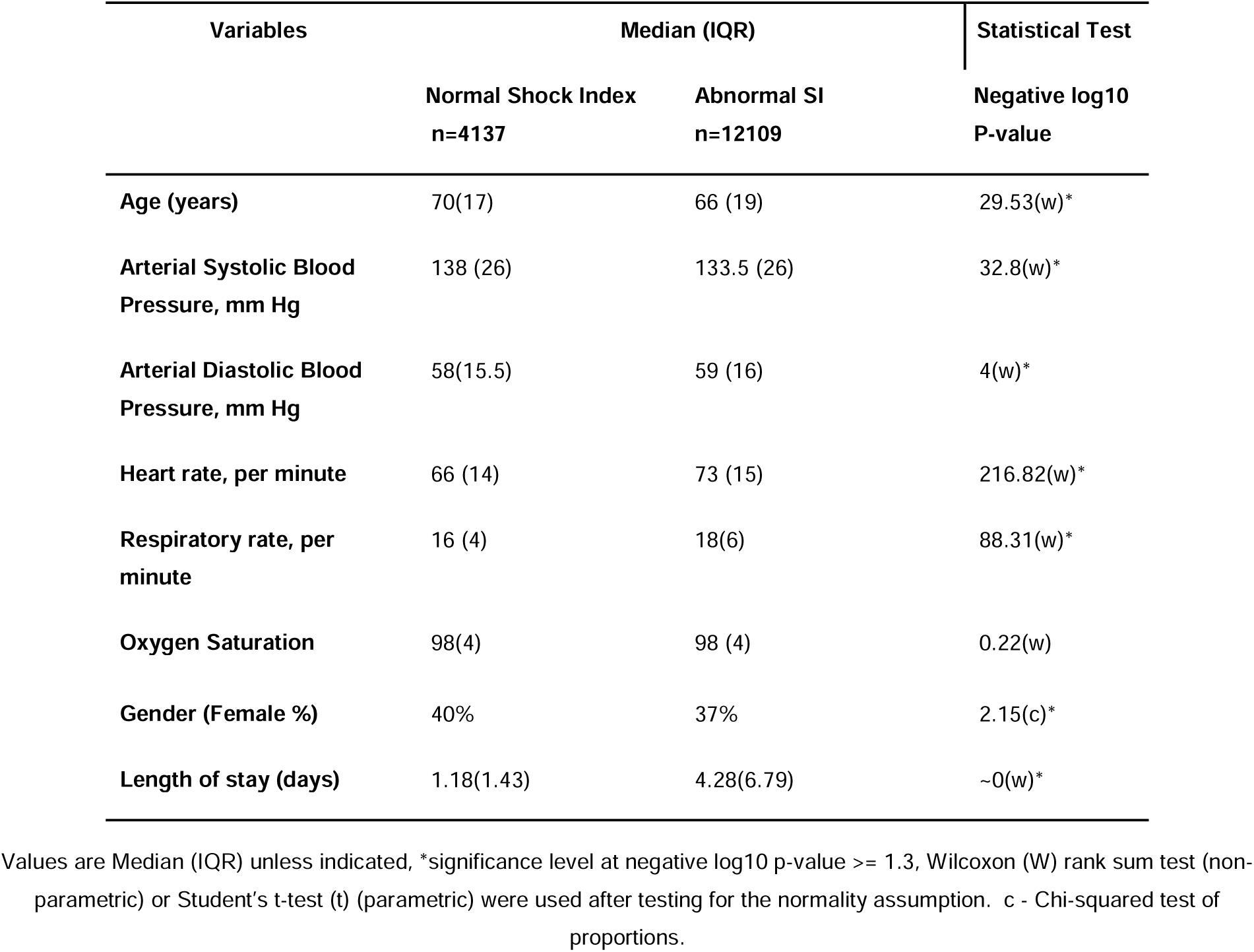
Characteristics of the eICU cohort observational window captured 4 hours earlier than the abnormal SI.

| Variables | Median (IQR) |  | Statistical Test |
| --- | --- | --- | --- |
|  | Normal Shock Index<br>n=4137 | Abnormal SI<br>n=12109 | Negative log10<br>P-value |
| Age (years) | 70(17) | 66 (19) | 29.53(w)* |
| Arterial Systolic Blood Pressure, mm Hg | 138 (26) | 133.5 (26) | 32.8(w)* |
| Arterial Diastolic Blood Pressure, mm Hg | 58(15.5) | 59 (16) | 4(w)* |
| Heart rate, per minute | 66 (14) | 73 (15) | 216.82(w)* |
| Respiratory rate, per minute | 16 (4) | 18(6) | 88.31(w)* |
| Oxygen Saturation | 98(4) | 98 (4) | 0.22(w) |
| Gender (Female %) | 40% | 37% | 2.15(c)* |
| Length of stay (days) | 1.18(1.43) | 4.28(6.79) | ~0(w)* |

Values are Median (IQR) unless indicated, \*significance level at negative log10 p-value $\geq 1.3$ , Wilcoxon (W) rank sum test (non-parametric) or Student's t-test (t) (parametric) were used after testing for the normality assumption. c - Chi-squared test of proportions.

Details of the included and excluded cohorts for the external validation sets, extracted from MIMIC-III and SafeICU, are shown in Supplemental Materials 3b and 3c. Data characteristics of external validation sets are shown in (Supplemental Materials 4, 5). SI status within the first 3 days of ICU stay was associated with both ICU mortality (all p values <0.05) and LoS (all p values <0.001) in all three datasets (Supplemental Materials 6,7). These associations suggest that early prediction of abnormal SI may help reduce adverse clinical outcomes, including mortality and prolonged length of stay (Supplemental Materials 6, 7).

### Model performance on eICU data

Deep learning models such as the CNN-LSTM model achieved a maximum percentage mean (SD) AUROC of 80(2), and a percentage mean (SD) AUPRC of 89(2). Bidirectional-LSTM achieved a maximum percentage mean (SD) AUROC of 83(1), and percentage mean (SD) AUPRC of 90(1). The TS-Feature set included 51 classes of 3970 linear and nonlinear time series features listed in (Supplemental Material 2). Of these, 2,120 were statistically significant for variable importance z-scores in the Boruta feature selection analysis. From the models trained on TS-features, GBM achieved higher performance, with a percentage mean (SD) AUROC of 86 (1.2) and a percentage AUPRC of 93 (1.1) (Figure 2a, 2b, Supplemental Material 8a, 8b). It identified 92% of all the shock events with an overall precision of 81% (Figure 2c).

**Figure 2:**
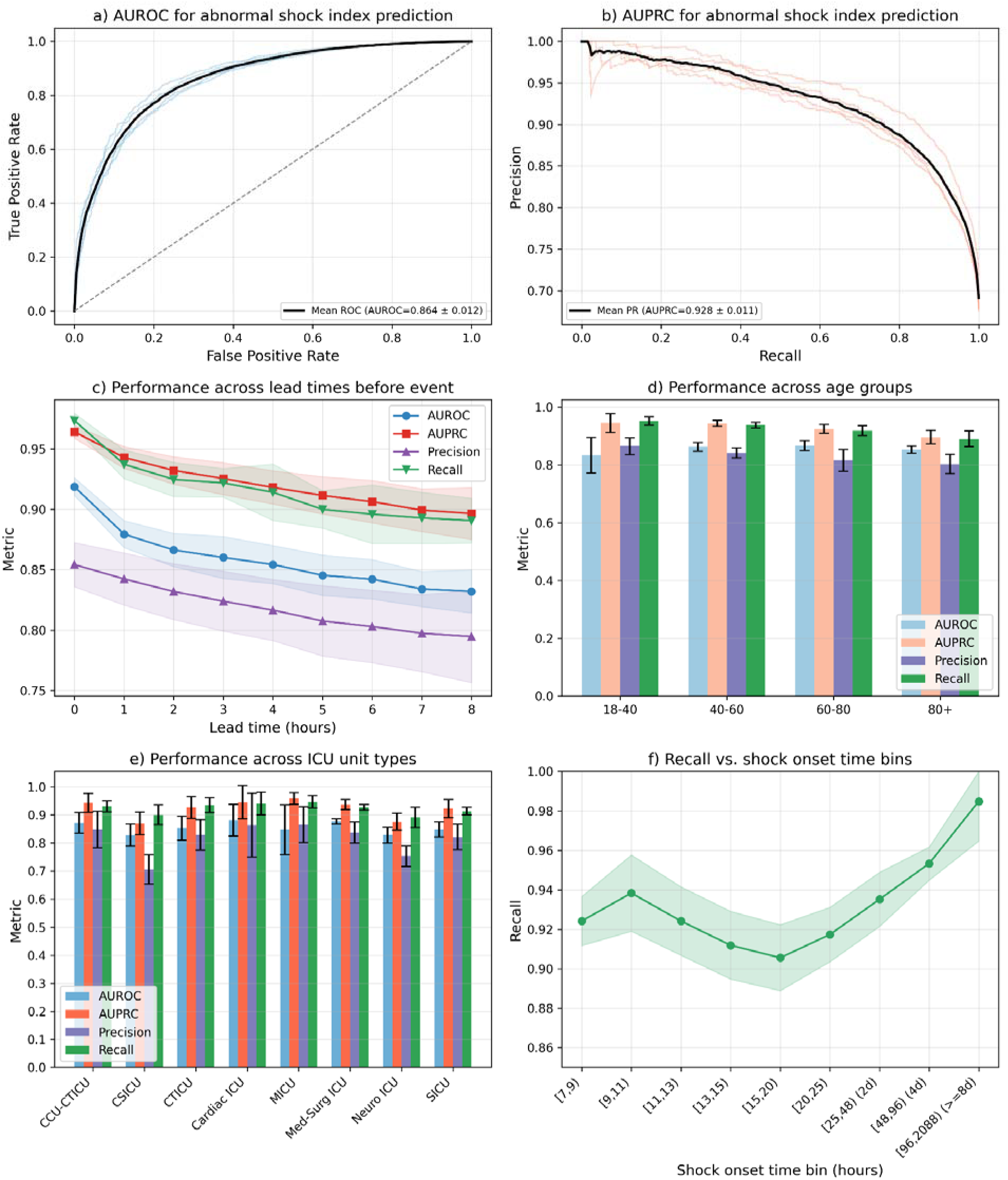
Model performance on eICU data. 2a) AUROC for the prediction of abnormal SI in the next 5 min to 8 hours. 2b) AUPRC for the prediction of abnormal SI in the next 5 min to 8 hours. 2c) Model performance for different lead times or times before the event. 2d) Model performance across different age groups: eICU data includes adult age groups. 2e) Model performance in different unit stay types (CCU – coronary care unit, CSICU- cardiac surgery ICU, CTICU-cardiothoracic ICU, MICU- medical ICU, SICU- surgical ICU). 2f) recall of the model for each time interval shown on the x-axis for the prediction made during different lengths of stays in the ICU.

The non-invasive variant of this model (SIgnose non-invasive) obtained a percentage mean (SD) AUROC of 81 (1), a percentage mean (SD) AUPRC of 90 (1), and an overall recall of 90% with a precision of 79%. Oversampling of the minority class and under-sampling of the majority class to correct for class imbalance did not meaningfully affect this model’s performance (Supplemental Material 9a, 9b). Based on these characteristics, the GBM model trained on TS-Features was advanced as the shock early warning system (SIgnose) for validation across ICUs.

### Model performance for different lead times, age groups, unit types, and onset time since admission

As with all the models tested, the performance of SIgnose decreased with increasing lead time from 0 to 8 hours. However, the model’s recall exceeded 89% for all lead times up to 8 hours (Figure 2c). The model performance indicators AUPRC and AUROC remained consistent across age groups, with a modest decline in AUPRC among older patients (above 60 years); AUROC remained stable in that age group, and recall exceeded 90% with precision exceeding 79% across all age groups (Figure 2d). All ICU types, except the cardiac surgery ICU and neuro ICU, had recall rates above 90%. The latter had marginally lower recall and precision, respectively (Figure 2e). The model’s overall recall remained well above 90% for all shock onset times (Figure 2f).

### Predictive Performance Analysis with Llama-3.1-8B model

To assess the potential of LLM-based embeddings, Llama-3.1-8B was applied to structured 5-minute-interval physiological time series data from eICU. Text prompts encoding demographic and physiological information were input into the model to extract contextual embeddings, which were then used in ML classifiers. In the eICU-only setup, models trained on eICU embeddings showed strong and consistent performance across classifiers (Figure 3.a), with SVM achieving the highest AUROC (0.88±0.01) and AUPRC (0.94±0.01) on the test set. A similar pattern was observed in the MIMIC-only evaluation (Figure 3b), where SVM again outperformed other models in terms of AUROC (0.84±0.02) and AUPRC (0.90±0.02), despite modestly lower overall performance compared to eICU. This result indicates that Llama-3.1-8B embeddings capture meaningful signals when training is siloed within the same dataset.

**Figure 3:**
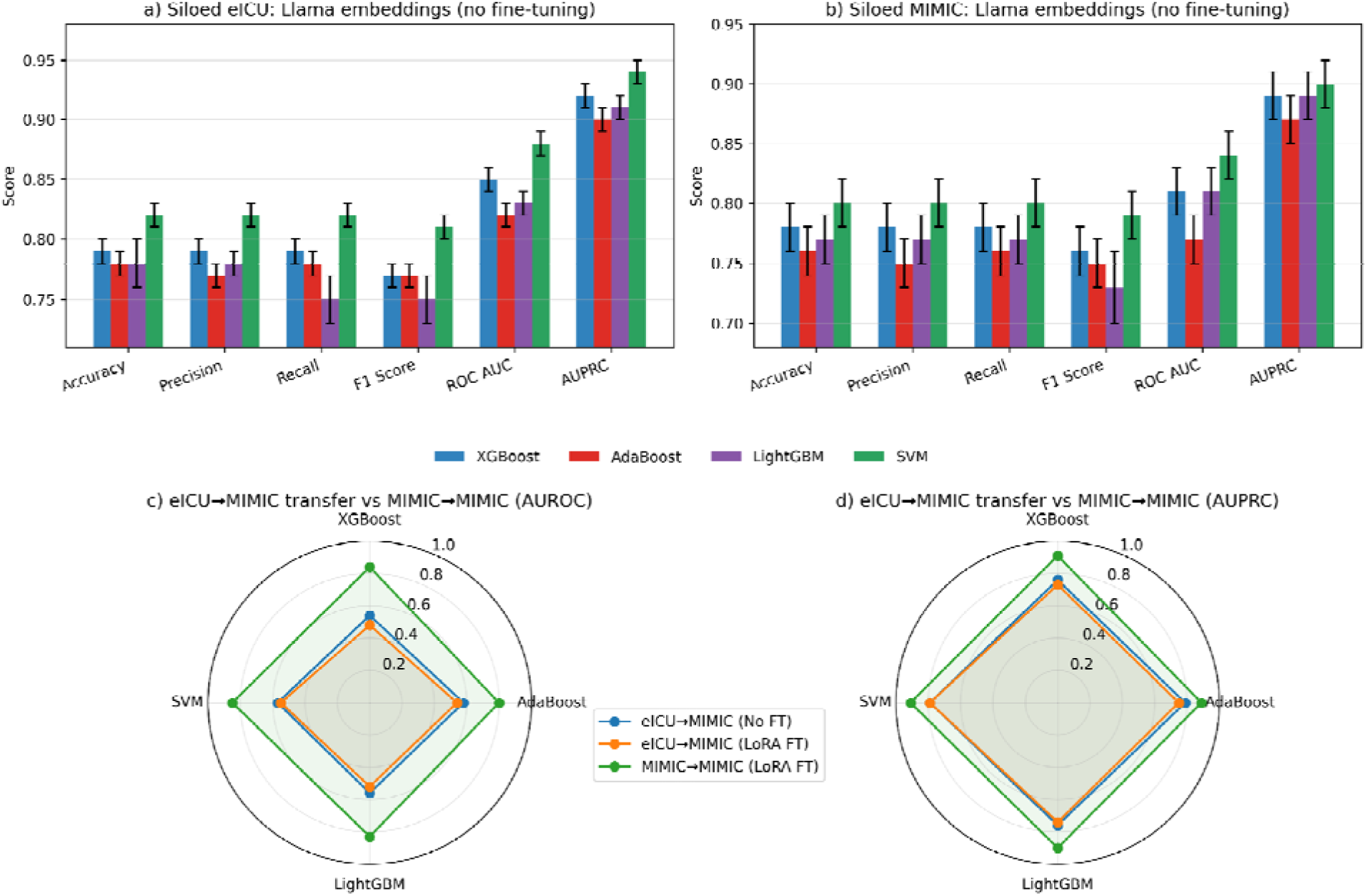
Performance of Llama⍰3.1⍰8B embeddings across tasks and transfer settings. (a) Siloed eICU results (no fine⍰tuning) for four classical classifiers across Accuracy, Precision, Recall, F1, AUROC, and AUPRC. (b) Siloed MIMIC results (no fine⍰tuning) with the same metrics. (c) AUROC for cross⍰cohort transfer from eICU→MIMIC with and without LoRA fine⍰tuning versus siloed MIMIC→MIMIC. (d) AUPRC for the same transfer settings, highlighting stronger within⍰dataset performance.

### Performance of external and prospective validation

We evaluated the robustness of Llama-3.1-8B embeddings under cross-dataset and siloed settings using eICU and MIMIC-III cohorts (Figure 3c, 3d). When models were trained on eICU and tested on MIMIC-III without fine-tuning, performance declined markedly across all classifiers, with AUROC values below 0.60 and AUPRC ranging from 0.76 to 0.79, indicating limited cross-dataset generalization. Parameter-efficient fine-tuning of Llama-3.1-8B using LoRA on eICU data did not substantially improve transfer performance, and AUROC values remained low across models. When the fine-tuned Llama-3.1-8B model was trained and evaluated within the MIMIC-III dataset, predictive performance was comparable to the non-fine-tuned setting, with a modest increase observed for SVM (AUROC: 0.85 ± 0.02; AUPRC: 0.91 ± 0.03). These results indicate that while Llama-3.1-8B embeddings are effective in siloed settings, their transferability across heterogeneous ICU cohorts remains limited.

In contrast, SIgnose, trained solely on eICU data, generalized better to both MIMIC-III and SafeICU cohorts without retraining. This underscores the strength of time-series-based modeling in capturing transferable patterns of physiological deterioration and its potential for deployment in varied ICU settings. Model generalization scenario of SIgnose with fine-tuning was significantly higher (p<0.0001, DeLong’s test) than scenarios without fine-tuning and a 2% increase over siloed models. This achieved an AUPRC of 93.5% (95% CI 93.0–94.0%) on MIMIC-III (Figure 4a) and 91.5% (95% CI 89.8–93.0%) on SafeICU (Figure 4b), and an AUROC of 87.0% (95% CI 86.3–87.6%) on MIMIC-III and 87.5% (95% CI 86.3– 88.8%) on SafeICU in five-fold cross-validation. Therefore, this approach demonstrated a strong model performance on external validation sites, as seen in (Figure 4c, 4d). Without fine-tuning, the direct application of the SIgnose on MIMIC-III and SafeICU data achieved an AUROC of 78% and 51%, respectively, thus demonstrating the need to contextualize the pre-trained models (Figure 4b, 4d). Furthermore, fine-tuning had a greater effect on precision, increasing it by up to 15%, compared with recall (Figure 4e, 4f). The performance of the fine-tuned model across different age groups, new ICU types, and time since admission is shown in Supplemental Material 11. Prospective validation of the fine-tuned model on the New Delhi’s SafeICU site yielded AUROC of 88.5% (95% CI 86.8–90.2%) and AUPRC of 91.1% (95% CI 88.2–93.6%) (Figure 5a, 5b), respectively, thereby validating its utility for abnormal SI prediction in this setting.

**Figure 4:**
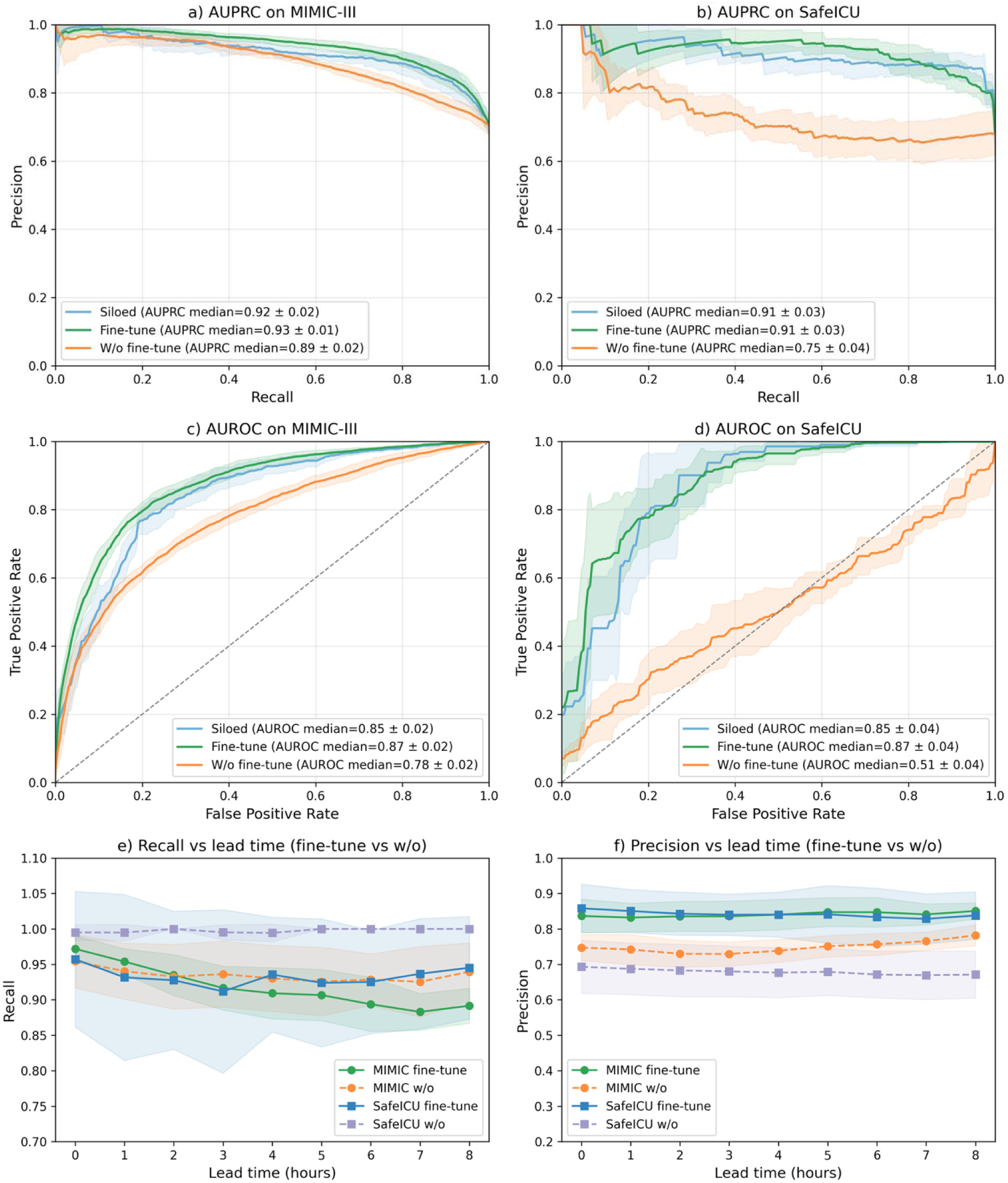
SIgnose Performance 4a, b) AUPRC on MIMIC-III and SafeICU data, respectively, for siloed, fine-tuning, and without fine-tuning. 4c, d) AUROC on MIMIC-III and SafeICU data, respectively, for siloed, fine-tuning, and without fine-tuning. 4e) Comparison of recall values for fine-tuned versus non-fine-tuned models on both the SafeICU and MIMIC-III datasets. 4f) The precision of fine-tuned and non-fine-tuned models on SafeICU and MIMIC-III data.

**Figure 5:**
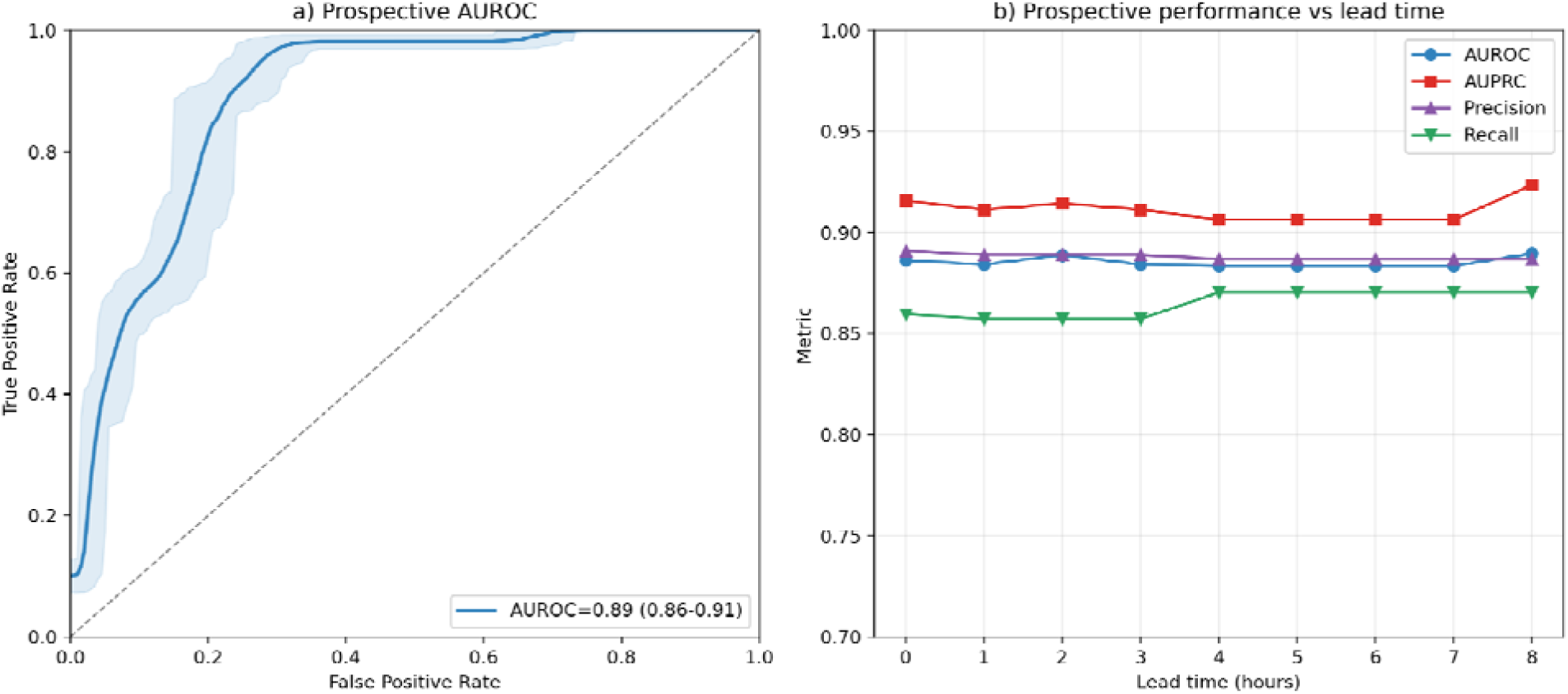
a) AUROC for the prospective validation of SafeICU data. b) Lead time-wise model performance indicators for the prospective validation of *SIgnose*.

### Model Interpretability Analysis

The *SIgnose* model is interpretable and clinically meaningful as it captures non-intuitive complexity features such as correlation dimension, recurrence quantification, and Benford correlations as important predictors of future development of abnormal SI, in addition to intuitive features such as minimum heart rate (Figure 6a, 6b, 6c, 6d). Importantly, even these intuitive features are not intended to perform the trivial task of phenotyping the patient, but rather to support the non-trivial future prediction of shock, as is commonplace in other complex prediction tasks, such as weather forecasting. The analysis highlighted vital signs such as heart rate, arterial blood pressure, respiratory rate, and patient age, along with derived statistical measures from these parameters, as primary contributors to the model’s output.

**Figure 6:**
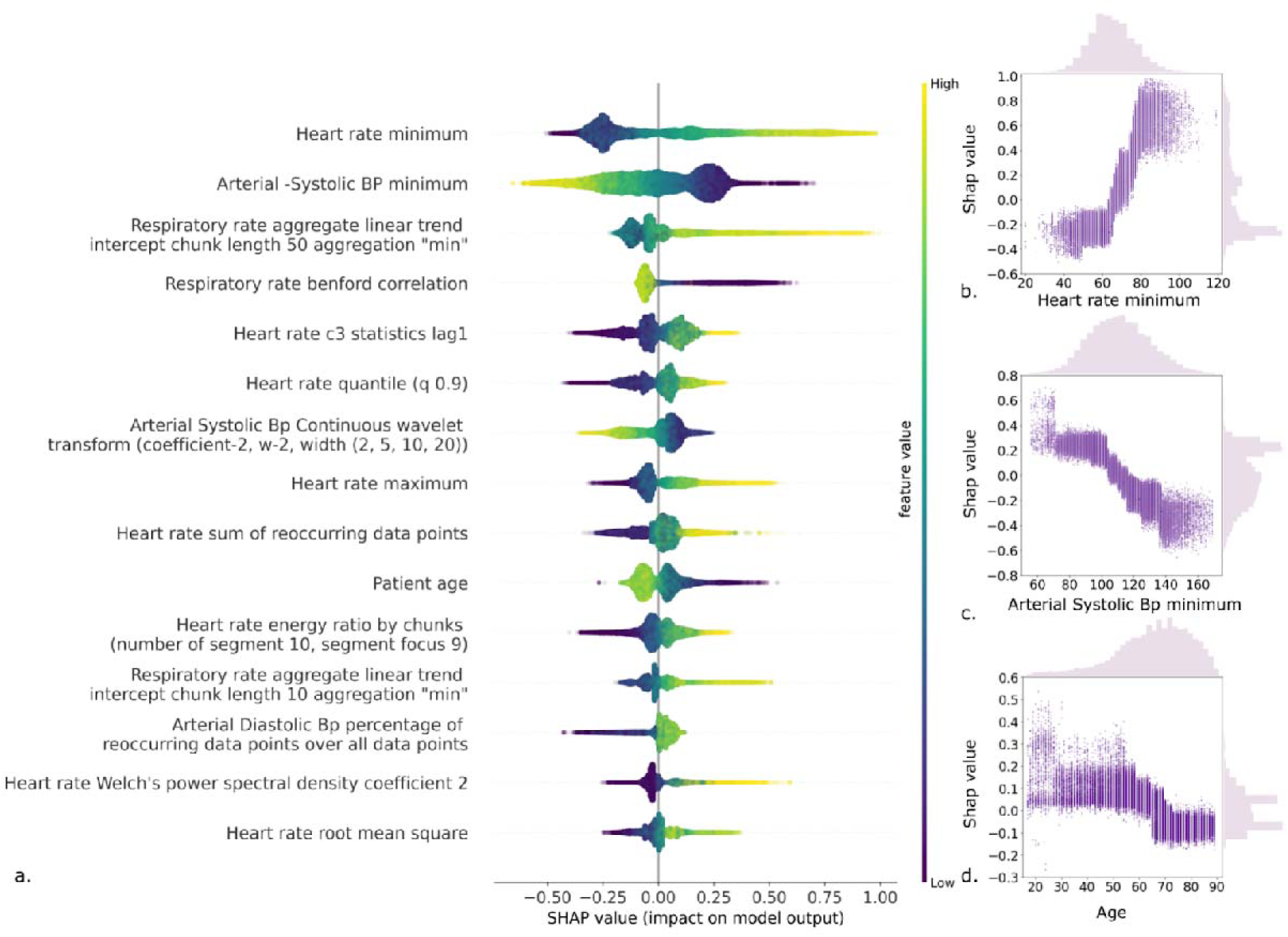
Model Interpretability Analysis. 6a) SHAP values of the top 15 TS-features (descending order) derived from the five-fold test set on eICU data. The Violin plot shows the distribution of SHAP values. The color map represents the feature value from low to high. 6b) A scatter plot of Heart rate minimum and corresponding SHAP values shows a positive correlation between the two. 6c) A scatter plot of Arterial Systolic Blood pressure minimum and corresponding SHAP values, showing a negative correlation. 6d) A scatter plot of patient age and corresponding SHAP values shows a negative correlation. In addition to easily interpretable features such as minimum heart rate, we find that complexity features, such as correlation dimension, recurrence quantification, and Benford correlations, are important predictors of future abnormal SI development.

Heart Rate Minimum emerged as the top feature influencing the model’s predictions. Lower heart rate values were associated with a decreased risk of hemodynamic shock. In contrast, higher minimum heart rates, approaching 100 beats per minute (bpm), were associated with a higher risk of hemodynami shock. Clinically, this suggests that elevated minimum heart rates may reflect sympathetic overactivity or reduced parasympathetic tone, indicating physiological stress that could precede hemodynamic deterioration.

Arterial Systolic Blood Pressure Minimum was an important predictor. Lower minimum systolic blood pressure values (e.g., below 100 mmHg) increased the risk of hemodynamic deterioration, while higher values reduced it. This underscores the clinical importance of maintaining adequate systolic blood pressure to ensure sufficient organ perfusion and prevent the onset of shock.

The Respiratory Rate Aggregate Linear Trend, particularly the intercept over chunk lengths of 50 with minimum aggregation, was identified as an essential feature. Marked negative trends in respiratory rate over time were associated with a higher predicted risk of hemodynamic deterioration. This indicates that a decreasing respiratory rate may signal respiratory fatigue or central depression, which can compromise oxygenation and contribute to hemodynamic compromise.

The Respiratory Rate Benford Correlation showed that deviations from expected respiratory rate patterns were associated with higher risk predictions. Clinically, irregular breathing patterns detected through this correlation may reflect underlying respiratory distress or metabolic abnormalities that can exacerbate hemodynamic deterioration.

Heart Rate c3 Statistics Lag1, representing short-term heart rate variability, was a key predictor. Lower lag1 values increased the model’s predicted risk. This suggests that diminished short-term variability may reflect impaired autonomic regulation, making patients more vulnerable to hemodynamic fluctuations.

The 90th Percentile of Heart Rate Values (quantile q0.9) was influential, with higher values associated with higher risk predictions. A high 90th percentile indicates episodes of tachycardia, which may signify compensatory mechanisms in response to stress but can lead to cardiac strain if sustained.

Arterial Systolic Blood Pressure Continuous Wavelet Transform Coefficients captured multi-scale variations in the signal. Higher coefficients corresponded to increased variability, associated with a higher predicted risk of instability. This reflects that marked fluctuations in systolic blood pressure may indicate vasomotor instability, a hallmark of progressing shock states.

Heart Rate Maximum and the sum of Recurring Heart Rate Data Points strongly influenced the model’s predictions. Higher maximum heart rates increased the predicted risk, while a greater sum of recurring data points was associated with lower risk. This suggests that extreme elevations in heart rate can be detrimental, whereas consistent heart rate readings may indicate cardiovascular stability.

Patient Age was a key predictor: older patients had higher SHAP values, indicating a higher risk of hemodynamic deterioration. This highlights that aging is associated with decreased physiological reserves and increased comorbidities, making elderly patients more susceptible to shock.

The Heart Rate Energy Ratio (Chunked Segments) was associated with higher risk predictions. Clinically, a higher energy ratio may indicate abnormal heart rate dynamics or arrhythmias, which can compromise cardiac output.

Respiratory Rate Aggregate Linear Trend over chunk lengths of 10, with minimum aggregation, showed that negative trends over shorter chunks were associated with a higher risk prediction. Short-term decreases in respiratory rate may reflect acute changes in respiratory function and necessitate prompt assessment to prevent respiratory failure.

The Percentage of Recurring Arterial Diastolic Blood Pressure Data Points increased the model’s predicted risk when high. A lack of variability in diastolic blood pressure may indicate diminished autonomic responsiveness or vascular stiffness, both of which can impair adequate perfusion.

Heart Rate Welch’s Power Spectral Density (PSD) Coefficients in specific frequency bands correlated with increased risk predictions. Alterations in PSD coefficients suggest changes in autonomic balance, potentially revealing underlying arrhythmias or cardiac dysfunction.

Heart Rate Root Mean Square (RMS) values were associated with risk; lower RMS values indicated higher predicted risk. A reduced RMS indicates decreased overall heart rate variability, suggesting impaired autonomic regulation and increased vulnerability to hemodynamic deterioration.

Samples that were misclassified (highlighted) vs correctly classified samples were shown in the SHAP decision plot in (Supplemental Material 10).

### Model validation across several diagnosis categories

SIgnose was evaluated on a range of disease-related outcomes in the MIMIC-III (adult setting) and SafeICU (pediatric setting) datasets (Supplemental Material 12). The MIMIC-III dataset showed the highest sensitivity (87%) and positive predictive value (PPV) (92%) across pneumonia, atrial fibrillation, and acute kidney injury. For patients with sepsis, SIgnose had the highest sensitivity and positive predictive value (PPV) in the pediatric cohort (93% and 100%, respectively), whereas, for patients with hypertension, it showed higher sensitivity and PPV (88% and 94%) than the MIMIC-III dataset hypertension (82% and 90%). This indicates that our model is broadly applicable to critical care illnesses.

## DISCUSSION

The development of ML-based early warning systems for ICUs has been an active area of research in recent years(9,18,34). However, many of these models have not been successfully implemented in clinical practice due to issues with generalizability, interpretability, and integration into clinical workflow(35). In this study, we developed a model called SIgnose, which uses routinely collected vital signs data to predict the onset of abnormal SI in critical care patients. The SI is a widely accepted objective indicator of a variety of outcomes in emergency and critical care settings. It can be easily calculated from routinely monitored vital signs data in real time, making it applicable to ICUs around the globe. Our results confirm the value of SI for predicting outcomes such as mortality and LoS across all three datasets.

To ensure clinical applicability, we focused on readily available vital signs data rather than more complex, heterogeneous, sparse EHR data, which can hinder the adoption of prediction models, particularly in conditions such as shock. The features used in the model were derived from time-series data on five physiological vital signs and selected for their availability at the bedside across a variety of settings, age groups, and geographic locations. Minimal preprocessing was performed to preserve physiological signals, and the choice of Kalman filtering was based on its established ability to impute missing values in physiological time series data (36,37). The winning model (GBM) was used as our final model for predicting abnormal SI due to its superior performance (Fig. 2a, 2b; Supplemental Material 8a, 8b). Recent research on tree-based models suggests that they exhibit distinct inductive biases that enable them to excel on tabular data (38). The model predicted 92% of all abnormal SI events 8 hours before onset, with similar performance on external validation datasets. An 8-hour prediction was chosen to provide reliable, sufficiently long actionable windows, thus balancing the need for early intervention with the need for accurate prediction. The model also performed well in a pediatric population, with an average area under the receiver operating characteristic curve (AUC) of 87% and area under the precision-recall curve (AUPRC) of 92% on an external validation dataset in India, surpassing the AUC of 82% reported in a previous study that used 36 variables, including vitals and laboratory investigations, to predict the onset of hemodynamic instability in pediatric patients(17). However, the study data differ, so they may not be directly comparable; nonetheless, this comparison is valuable because it allows us to assess the generalizability and accuracy of the SIgnose model compared to the available model for the pediatric population.

We also provided some computational guidelines for building clinically relevant predictive models, including the use of transfer learning to improve generalizability. In this study, we demonstrated the effectiveness of “feature transfer,” in which model features were transferred to different settings and fine-tuned to enhance overall AUC and precision while maintaining a high recall rate. We externally validated our model on the SafeICU pediatric dataset. We chose to evaluate our model on a pediatric cohort for several reasons. First, the physiological characteristics of the SI may differ between pediatric and adult patients, due to differences in body size, composition, and function. Second, transferring features from the adult to the pediatric setting demonstrates the generalizability of the approach across age groups and geographic settings.

Our model initially achieved an AUROC of 51% and an AUPRC of 75% on the SafeICU dataset, outperforming random chance and performing exceptionally well on imbalanced datasets, as evidenced by strong sensitivity, precision (PPV), and F1 score metrics (39). However, when we retrained the model on the SafeICU data, we observed a substantial performance improvement, with an AUROC of 88.5%. This suggests that our model can adapt to new data and generalize to different patient populations and care settings. Additionally, using SHAP analysis, we find that easily interpretable features, such as minimum heart rate, and complexity features, such as correlation dimension, recurrence quantification, and Benford correlations, are significant predictors of future abnormal SI development.

The model was also found to have broad applicability across different critical care illnesses, with particularly high sensitivity and positive predictive value for sepsis in the pediatric population and for pneumonia, atrial fibrillation, and acute kidney injury in the MIMIC-III dataset. For all patients at risk of shock, physiological hemodynamic deterioration is a final common pathway in many illnesses. We intended the model to be used for the early detection of this cascade that leads to multiple organ failure. We have profiled our model across the most frequent diagnoses and show that its performance is similar across conditions (Supplemental Material 12). A key strength of SIgnose is its performance despite the dynamic nature of SI regulation. While this confirms the presence of early warning signals before the initiation of therapeutic interventions or overt physiological decompensation, it also highlights the scope for future model improvement, including the inclusion of real-time interventions to improve predictions.

In parallel with the development of SIgnose, we evaluated the use of Llama-3.1-8B derived embeddings combined with classical machine learning classifiers for early prediction of abnormal SI across ICU datasets. We chose the Llama-3.1-8B model because it offers a strong balance between context length and computational efficiency, making it well-suited to resource-limited clinical settings. Its open-weight design enables reproducible and scalable hemodynamic prediction without dependence on proprietary models. Llama-3.1-8B embeddings yielded strong predictive performance when training and evaluation were conducted within the same dataset, with SVM achieving AUROC values of 0.88 on eICU and 0.84 on MIMIC-III and showing a modest improvement on MIMIC-III after fine-tuning (AUROC: 0.85). However, when models learned from eICU were transferred to MIMIC-III, predictive performance declined substantially across classifiers, and fine-tuning on the source dataset did not meaningfully mitigate this degradation. These findings indicate that while large foundation models offer powerful representation learning capabilities, their computational cost, hardware dependence, and limited transportability across heterogeneous clinical environments pose challenges for bedside deployment. In contrast, SIgnose demonstrates that physiologically grounded, feature-based models can achieve strong multicenter generalization with substantially lower computational overhead.

The SHAP analysis provided valuable insights into the features influencing the risk prediction of hemodynamic shock. The identification of minimum heart rate as the top predictor aligns with clinical observations that elevated minimum heart rates may reflect decreased heart rate variability and increased sympathetic activity, both of which are linked to adverse outcomes in critically ill patients. Reduced heart rate variability is a known predictor of mortality in sepsis patients, emphasizing the importance of monitoring this parameter.

The significance of the arterial systolic blood pressure minimum as a predictor underscores the clinical relevance of blood pressure monitoring. Low systolic blood pressure indicates inadequate cardiac output and tissue perfusion, often preceding shock. Continuous monitoring allows for early detection and intervention (Supplemental Material 2, Row 9).

The findings related to respiratory rate trends highlight the importance of respiratory monitoring. Negative trends in respiratory rate may indicate respiratory fatigue or impending failure, which often precedes cardiovascular collapse in patients developing shock. Early detection facilitates timely interventions to prevent deterioration (Supplemental Material 2, Row 13).

The association of Benford correlation in respiratory rate with increased risk suggests that deviations from expected respiratory patterns may indicate irregular breathing, common in respiratory distress or metabolic dysfunction associated with shock. Utilizing statistical methods such as Benford’s Law can enhance the detection of such anomalies (Supplemental Material 2, Row 16).

The identification of heart rate variability measures, such as c3 statistics, lag1, and RMS, as key predictors align with the understanding that reduced heart rate variability is associated with autonomic dysfunction and poor prognosis in critically ill patients. Monitoring these parameters can aid in detecting early signs of deterioration (Supplemental Material 2, Row 6).

The influence of the 90th percentile of heart rate values indicates that sustained tachycardia can signal compensatory mechanisms under stress, potentially leading to cardiovascular exhaustion if prolonged. Recognizing and managing sustained tachycardia is crucial in patient care (Supplemental Material 2, Row 5).

The use of advanced analytical techniques, such as the continuous wavelet transform applied to arterial systolic blood pressure, enables the detection of subtle hemodynamic changes that serve as early indicators of shock. This method enhances the clinician’s ability to identify and respond to instability promptly (Supplemental Material 2, Row 2).

The impact of the maximum heart rate and the sum of recurring data points on risk prediction highlights the balance between physiological stress and stability. Elevated maximum heart rates may signal excessive stress or compensatory mechanisms, while stable heart rates reflect a controlled cardiovascular state (Supplemental Material 2, Rows 7 and 17).

The role of patient age as a predictor emphasizes the increased vulnerability of older patients due to reduced physiological reserve and comorbidities. This finding supports the need for heightened vigilance and tailored interventions in older patient populations (Supplemental Material 2, Row 4).

The significance of the heart rate energy ratio and Welch’s PSD coefficients points to the importance of analyzing heart rate signals for abnormal variability or arrhythmias. These analytical approaches can enhance early detection and management of arrhythmias, improving patient outcomes (Supplemental Material 2, Row 10).

Lastly, the relevance of variability in arterial diastolic blood pressure and short-term respiratory trends to risk prediction underscores the multifaceted nature of hemodynamic monitoring. Recognizing patterns indicative of poor vascular tone or respiratory fatigue allows for timely interventions to improve vascular tone, perfusion, and respiratory function (Supplemental Material 2, Rows 13 and 14).

In summary, the SHAP analysis elucidated the key physiological parameters influencing hemodynamic shock risk predictions. Understanding the clinical implications of these features enables healthcare professionals to focus on monitoring and managing the most impactful parameters. This targeted approach has the potential to improve patient outcomes through early detection and timely intervention.

Several limitations warrant consideration. First, the primary SIgnose model requires invasive arterial blood pressure (ABP), which reduces cohort size and may limit generalizability to settings without routine ABP monitoring; a non-invasive variant was therefore evaluated, albeit with potentially reduced hemodynamic resolution. Second, abnormal SI is influenced by vasoactive therapy. Some patients who later develop abnormal SI may appear hemodynamically stable during the observation window due to treatment-related compensation, and the model may capture early signatures of impending loss of this compensation rather than untreated shock alone. To explore this, we performed an ablation excluding systolic and diastolic blood pressure features, with preserved predictive signal suggesting complementary information from heart rate variability, respiratory dynamics, and nonlinear features; these findings are hypothesis-generating. Future work will focus on treatment-aware stratification. Leveraging existing treatment charts alongside time-series features and embedding pipelines will enable systematic evaluation of how vasoactive therapy modulates physiologic variability and model performance. Third, as with any early-warning system, alarm fatigue is a concern. Predictions were generated at hourly intervals to limit alert burden, but safe deployment will require careful integration with clinical workflows (40). Finally, SI does not capture all shock phenotypes (e.g., bradycardia-mediated shock). Future work will focus on treatment-aware stratification using medication data and on adaptive SI thresholds to better reflect shock severity and evaluate effects on patient-centered outcomes.

In this work, we present SIgnose, a monitor-only early-warning framework for identifying impending abnormal SI from continuously streaming ICU vital signs. Across multicenter development, external validation, and prospective pediatric ICU evaluation, SIgnose demonstrated robust generalization using physiologic variability and nonlinear time-series features derived from routine bedside monitoring, without reliance on laboratory data, clinician charting, or computationally intensive foundation-model inference. Our comparison with Llama-3.1-8B embeddings highlights that while foundation-model representations can perform well in siloed settings, their cross-site generalization and computational demands may limit feasibility for continuous bedside deployment, particularly in time-sensitive critical care settings. Beyond predictive performance, the model highlights clinically plausible early signatures of hemodynamic deterioration, including changes in heart rate variability, respiratory dynamics, and multiscale blood pressure patterns that may precede overt instability or loss of treatment compensation. In an era increasingly dominated by GPU-intensive AI, SIgnose underscores the value of low-cost, interpretable, and real-time bedside models that are deployable across diverse critical care environments. We have made it openly available to facilitate its use and further development. Instructions for installing and operating the Docker container app are provided in Supplemental Material 14.

## Data Availability

All data produced in the present study are available upon reasonable request to the authors.

## List of abbreviations

AIIMS: All India Institute of Medical Science
AUPRC: Area Under the Precision-Recall Curve
AUROC: Area Under the Receiver Operating Characteristic Curve
BiLSTM: Bidirectional Long Short-Term Memory
CCU: Coronary Care
CNN LSTM: Convolutional Neural Network Long Short-Term Memory
CSICU: Cardiac Surgery
CTICU: Cardiothoracic
DL: Deep Learning
ICU: Intensive Care Unit
LLM: Large Language Model
LoS: Length of Stay
LSTM: Long Short-Term Memory
MICU: Medical Intensive Care Unit
MIMIC: Medical Information Mart for Intensive Care
ML: Machine Learning
PSD: Power Spectral Density
RMS: Root Mean Square
SI: Shock Index
SICU: Surgical Intensive Care Unit
SVM: Support Vector Machine
TS-features: Time-Series Features

## Declarations

### i) Funding

This work was supported by the Wellcome Trust/DBT India Alliance Fellowship IA/CPHE/14/1/501504 awarded to Prof.Tavpritesh Sethi. A CSIR-GATE fellowship supports Dr.Aditya Nagori. Mr. Pradeep Singh is supported by the Indo-Israel collaborative research grant awarded to Prof. Tavpritesh Sethi and Prof. Rakesh Lodha. Indian Council of Medical Research: AI-Adhoc-18/2022-AI Cell.

### ii) Conflicts of interest

All authors declare no financial or non-financial competing interests.

### iii) Ethics approval

- The study was approved by the Institutional Ethics Committee (IEC) of All India Institute of Medical Science (AIIMS), New Delhi, India, for new proposal (NP) reference number IEC/NP-211/08.05.2015 and IEC-787/07.10.2022, Revised Proposal (RP), RP-14/2022 and involved no change in routine patient care.
- The EQUATOR (Enhancing the QUAlity and Transparency Of Health Research) Network’s TRIPOD (31) guidelines were followed for developing the machine learning models and reporting results.

### iv) Consent to participate

Ethics approval was obtained for this study, and the requirement for individual consent was waived because all data were anonymized before use.

### v) Consent for publication

Not applicable

### vi) Code availability

The code underlying this study is freely available at https://github.com/tavlab-iiitd/SIgnose.

### vii) Availability of data and materials

The datasets used and analyzed during the current study are available from the corresponding author upon reasonable request. The in-house clinical data are not publicly available due to ethical and privacy considerations aimed at protecting patient confidentiality. The trained models, implementation details, and feature generation pipelines used in this study are publicly available at: https://github.com/tavlab-iiitd/SIgnose.

### viii) Authors’ contributions

AN and TS designed the study. AN, PS, TS, and RL were involved in acquiring data. AN and PS performed pre-processing and developed the cohorts. AN, PS, SF, and AG were involved in feature computation and feature selection. AN, SF, PS, AG developed models and performed model generalization, AN, VV formal analysis, and visualized the results. AN, VV, and RA were involved in statistical analysis. PS, AN, AD, NN, and AM assessed LLM-based predictive modelling; and AN, TS, PS, and AD interpreted the data and wrote the first draft of the report. TS, AN, and RL critically revised the report for its important intellectual content. HB, PA, PS, and AN are involved in the development of Docker App. All authors read and approved the final version of the manuscript and agree to be accountable for all aspects of the work.

## ix) Acknowledgements

Authors acknowledge support from the Wellcome Trust/DBT India Alliance Fellowship: IA/CPHE/14/1/501504, Indian Council of Medical Research: AI-Adhoc-18/2022-AI Cell, and also the Centre of Excellence in Healthcare, IIIT-Delhi. Also, thank Mr. Varun Prakash and Mr. Anil Sharma for the technical support provided at the PICU at AIIMS, New Delhi.

## Notes

### Competing Interest Statement

The authors have declared no competing interest.

### Author Declarations

The study was approved by the Institutional Ethics Committee (IEC) of All India Institute of Medical Science (AIIMS), New Delhi, India, for new proposal (NP) reference number IEC/NP-211/08.05.2015 and IEC-787/07.10.2022, Revised Proposal (RP), RP-14/2022 and involved no change in routine patient care.

