## Supplemental Material for "Development and Prospective Validation of Predictive Model for Early Hemodynamic Deterioration in Critical Care: A Multicenter Study"

Supplemental Material 1: Physiological time-series data is labeled for each 30-minute time window as shock-positive or shock-negative. A 420-minute observation window is used to predict shock status up to 8 hours ahead.

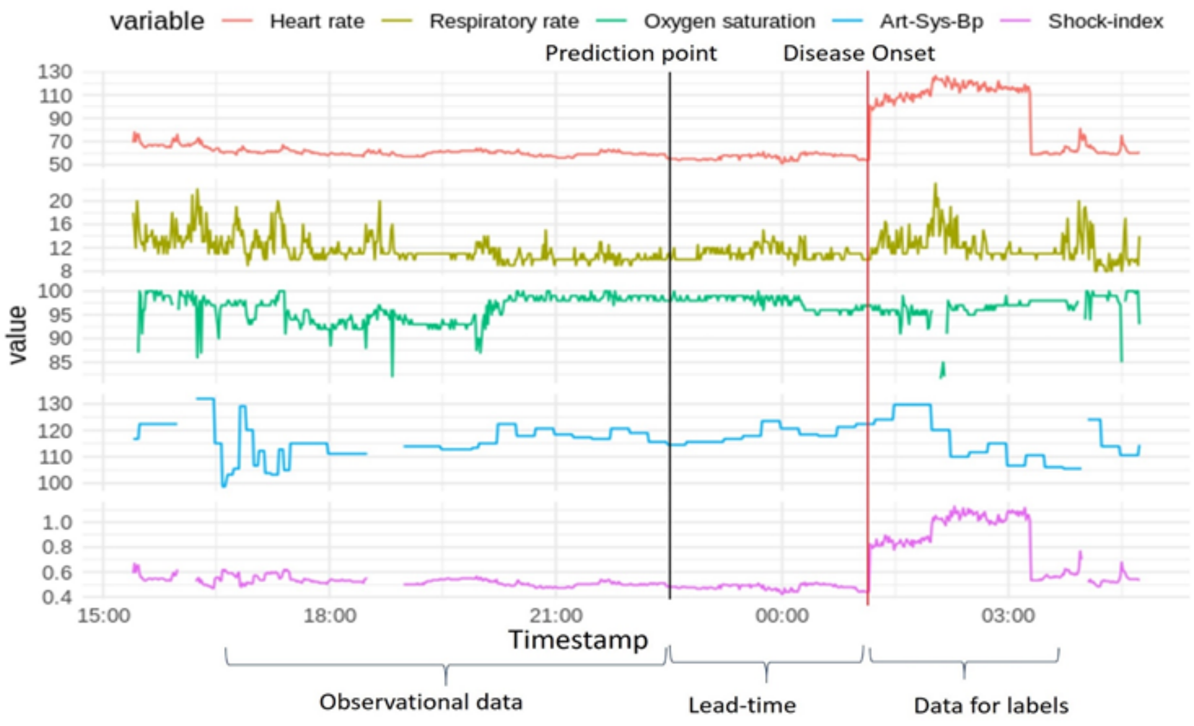


**Supplemental Material 1: Each 30-minute time window is labeled as shock-positive or shock-negative. A 420-minute observation window is used to predict shock status up to 8 hours ahead.**

**Supplemental Material 2:** Non-linear time-series features description.

| Sl.No | Feature | Description |
|---|---|---|
| **1** | Absolute Energy (abs_energy) | Returns the absolute energy of the time series. |
| **2** | Continuous Wavelet Transform Coefficients (cwt_coefficients) | CWT proposes a time scale illustration of a signal. The duration of the examined signal will help dynamically identify non-linearities. |
| **3** | Fast Fourier Transformation Coefficient (fft_coefficient) | This feature uses the Fourier transform algorithm to compute the Fourier coefficients of the one-dimensional discrete Fourier transform. |
| **4** | Mean | This feature will return the mean of x. |
| **5** | Quantile | Calculates the q quantile of x. The quantile divides the sample into equal-sized adjacent subgroups. |
| **6** | Lag (c3) | For all the time series, perfect correlation will be at lag=0. The shift in the time series will decrease the correlation value. |
| **7** | Sum of re-occurring data points (sum_of_reoccurring_datapoints) | Returns the sum of all the time series data points that occur more than once. |
| **8** | Sum (sum_values) | Will calculate the sum over the time series values. |
| **9** | Minimum | The minimum number is the least value among the given set of values. |
| **10** | Energy ratio by chunks (energy_ratio_by_chunks) | This is expressed as a ratio of the sum of all the squares of chunk i over the whole series of chunks. |
| **11** | Partial Autocorrelation (Partial_autocorrelation) | This provides the partial correlation for a time series under stationary conditions. |
| **12** | Aggregated linear trend (Agg_linear_trend) | This calculates the value for an aggregation function of a linear least squares regression. |
| **13** | Linear trend (linear_trend) | This will perform linear least-squares regression on one less than the length of the time series. |
| **14** | Asymmetric statistic (time_reversal_asymmetry_statistic) | This means that entropy will increase over time. |
| **15** | Approximate entropy (approximate_entropy) | It calculates the non-stationary and chaotic time series. |
| **16** | Binned entropy (binned_entropy) | It will first bin the values and then compute the entropy. |

| | | |
|---|---|---|
| **17** | Maximum | This feature calculates the highest among the time series values. |
| **18** | Mean change (mean_change) | It provides the mean value of the difference between time series values. |
| **19** | Median | This feature computes the median over time-series values. |
| **20** | Change quantiles (change_quantiles) | First, select the corridor on the y-axis, and then within this corridor, calculate the mean of the absolute change in the series. |
| **21** | Ratio beyond sigma (ratio_beyond_r_sigma) | These are the ratios beyond the r-sigma and are away from the series mean. |
| **22** | Index mass quantile (index_mass_quantile) | These calculate the relative index at which x% of the mass of the time series lies to the left. |
| **23** | Sum of reoccurring values (sum_of_reoccurring_values) | These are the sum of all the repeated values in the series. |
| **24** | Longest strike below mean (longest_strike_below_mean) | This is the length of the longest strike (consecutive subsequence) which is below the mean of the time series. |
| **25** | Continuous Wavelet Transform peak (number_cwt_peaks) | After smoothing the time series with a Ricker wavelet, this feature returns the number of peaks. |
| **26** | Autocorrelation | This provides the partial correlation for a time series under a stationary condition. |
| **27** | Largest fixed point of the dynamics (max_langevin_fixed_point) | When the time series is fitted to the Langevin model (with deterministic dynamics), this parameter gives the largest fixed point of the dynamics. |
| **28** | percentage_of_reoccurring_values_to_all_values | In the given time series, this parameter gives the normalized ratio of all unique values that appear more than once. |
| **29** | ratio_value_number_to_time_series_length | This measure is closely related to percentage_of_reoccurring_values_to_all_values, as this gives an output equal to 1 if every single value in the time series occurs only once. If the case isn't so, the output value would be less than 1. |
| **30** | Standard_deviation | Calculates the standard deviation of the time series. Expresses a measure that signifies how much all the |

| | | |
|---|---|---|
| | | values of the time series differ from the mean value of the time series. |
| **31** | Variance | Calculates the variance of the time series. It is the square of the standard deviation. |
| **32** | Autoregressive coefficient (ar_coefficient) | For a given time series, first fit an autoregressive process's unconditional maximum likelihood to it, then calculate the AR coefficients. |
| **33** | sample_entropy | This parameter provides us with a measure of complexity. Closely related to approximate entropy, it has several advantages over it, such as data length independence and lower computational cost. |
| **34** | fft_aggregated | This feature contains several properties of the time series, such as skew and variance. |
| **35** | kurtosis | In a frequency distribution curve of a time series, this parameter measures how sharp the peak is. |
| **36** | Percentage_of_reoccurring_datapoints_to_all_datapoints | In a given time series, this parameter gives the percentage of unique values that appear more than once. This is different from Percentage_of_reoccurring_values_to_all_values, as this percentage is normalized with respect to the unique values, rather than all of the data points. |
| **37** | skewness | Skewness measures the asymmetry in the time series, or, in other words, shows on which side the time series leans (left or right). |
| **38** | Cid_ce : | This function calculator estimates the complexity of a time series. |
| **39** | Last_location_of_minimum : | The location of the minimum value of x I'd calculated with respect to the length of x. |
| **40** | Spkt_welch_density | In this feature calculator, the first step is to shift the time series x from the time domain to the frequency domain. Then it estimates the cross-power spectral density. |
| **41** | Count_below_mean | This function returns the number of x-values that may fall below the mean of x. |
| **42** | agg_autocorrelation | This function takes different lags into account and then calculates the value of an (fgg) aggregation function over the (R(l)) autocorrelation. |

| | | |
|---|---|---|
| **43** | Number_peaks : | In time series x, this function will calculate the number of peaks that are at least n. |
| **44** | Absolute_sum_of_changes: | This function returns the sum of the absolute value of the consecutive changes in the time series x. |
| **45** | Mean_abs_change | This function returns the mean of the absolute differences between successive values in the time series. |
| **46** | Longest_strike_above_mean: | This function returns the length of the longest consecutive subsequence in x that is bigger than the mean of x. |
| **47** | Friedrich_coefficients: | It is the coefficient of the polynomial h(X), where h(x) is fitted to the deterministic dynamics of the Langevin model. |
| **48** | Augmented_dickey_fuller: | The test, which checks whether a time series sample contains a unit root, is called the augmented Dickey-Fuller test and is a hypothesis test. |
| **49** | First_location_of_maximum: | The first location of the maximum value of x is calculated with respect to the length of x. |
| **50** | First_location_of_minimum | The first location of the minimum value of x is calculated with respect to the length of x. |
| **51** | Mean_second_derivative_central: | This function will return the mean value for the central approximation of the second derivative. |

**Supplemental Material 3:** Data preparation and cohort formation.

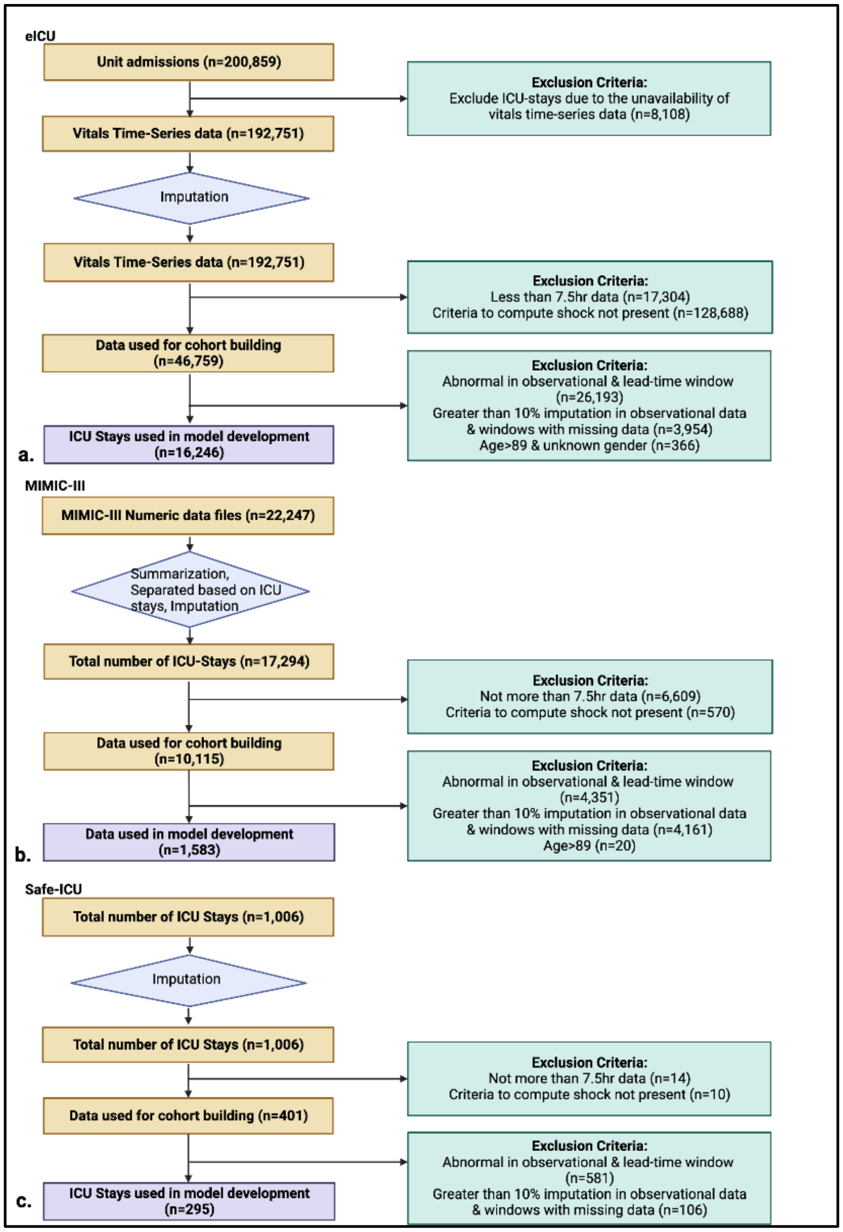


**Supplemental Material 3: Data Preparation and Cohort Formation 3a.) In eICU, 200,859 admissions were used as the starting point, with 8,108 ICU-stays removed due to the unavailability of the physiological time series data 3b.) MIMIC-III and 3c.) SafeICU pediatric cohort formation.**

**Supplemental Material 4:** Characteristics of the MIMIC cohort.

| Variables | Median (IQR) | | Statistical Test |
|---|---|---|---|
| | **Normal Shock Index n=482** | **Abnormal Shock Index n=1101** | **Negative log10 P-value** |
| **Age (years)** | 65.76(21.06) | 65.57(20.04) | 0.057(w)* |
| **Arterial Diastolic Blood Pressure, mm Hg** | 60.4(6.4) | 59.1(5.64) | 0.471(w)* |
| **Arterial Systolic Blood Pressure, mm Hg** | 129(11.51) | 118.45(10.6) | 16.03(w)* |
| **Heart rate, per minute** | 69(4.43) | 86.35(4.71) | 84.61(w)* |
| **Respiratory rate, per minute** | 17.2(2.64) | 18(2.9) | 4.48(w)* |
| **Oxygen Saturation** | 97(1.17) | 97(1.35) | 0.033(w)* |
| **Gender (Female %)** | 43% | 40% | 0.31(c)* |
| **Length of stay (days)** | 2.02(2.82) | 3.56(4.89) | ~0(w)* |

**Supplemental Material 5:** Characteristics of the SafeICU pediatric cohort.

| Variables | Median (IQR) | | Statistical Test |
|---|---|---|---|
| | **Normal Shock Index n=89** | **Abnormal Shock Index n=206** | **Negative log10 P-value** |
| **Age (months)** | 18.5(80.09) | 5.12(15.18) | 6.97(w)* |
| **Arterial Diastolic Blood Pressure, mm Hg** | 50.06(3.5) | 48.68(0.58) | 1.68(w)* |
| **Arterial Systolic Blood Pressure, mm Hg** | 94.96(5.01) | 93.06(0.96) | 1.01(w)* |
| **Heart rate, per minute** | 132.85(8.14) | 128.5(7.12) | 1.53(w)* |
| **Respiratory rate, per minute** | 31(4.53) | 34.98(4.38) | 2.33(w)* |
| **Oxygen Saturation** | 97(2.32) | 95.85(2.43) | 4.44(w)* |
| **Gender (Female %)** | 37% | 38% | 0.03(c) |
| **Length of stay (days)** | 4.16(5.20) | 9.63(11.98) | 196.94(w)* |

**Supplemental Material 6:** Association of mortality with the first three days of shock index status.

| Outcome | Cohort | Odds ratio | 95%CI | P-value |
|---|---|---|---|---|
| | eICU | 12.01 | 8.96-16.56 | <0.0001 |
| | MIMIC-III | 1.79 | 1.07-3.03 | 0.029756 |
| **Mortality** | SafeICU | 3.26 | 1.27-10.26 | 0.021693 |

**Supplemental Material 7:** Association of LOS with the first three days of shock index status.

| Outcome | Cohort | Regression Coefficient | 95%CI | P-value |
|---|---|---|---|---|
| | eICU | 4.64 | 4.41-4.88 | <2.2x10-6 |
| **LOS** | MIMIC-III | 1.20 | 0.79-1.61 | <0.0001 |
| | SafeICU | 4.69 | 2.19-7.17 | 0.0002 |

**Supplemental Material 8:** Model training on TS-features.

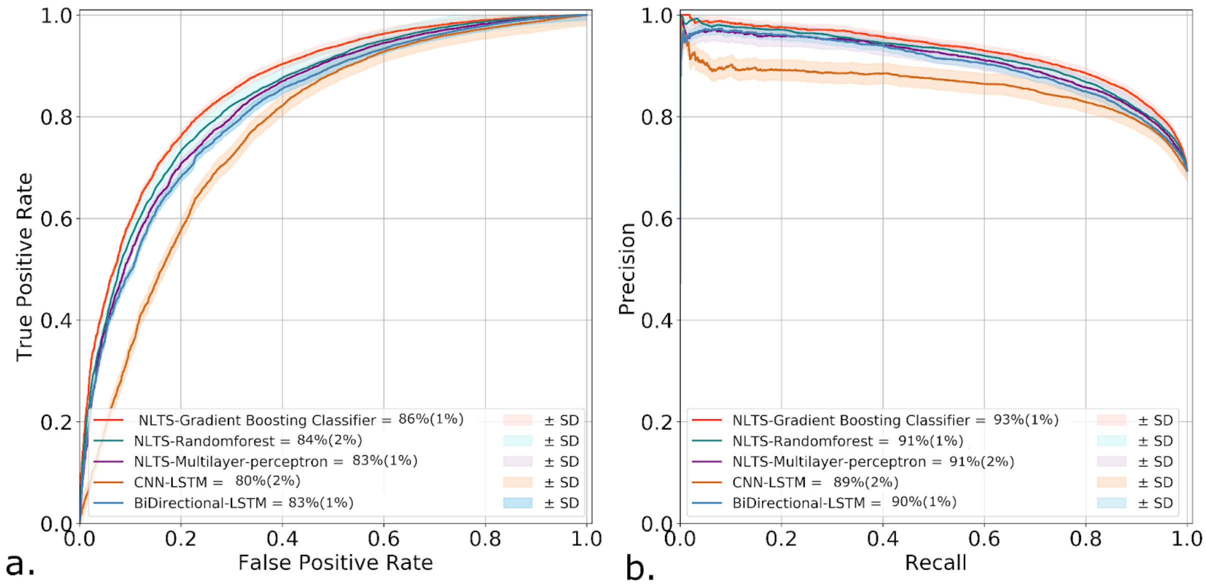


**Supplemental Material 8: 8a.) AUROC and 8b.) AUPRC for different models trained on the eICU hospital splits. The gradient boosting classifier was the best-performing model.**

**Supplemental Material 9:** *SIgnose* with oversampling of the minority class, and random undersampling of the majority class.

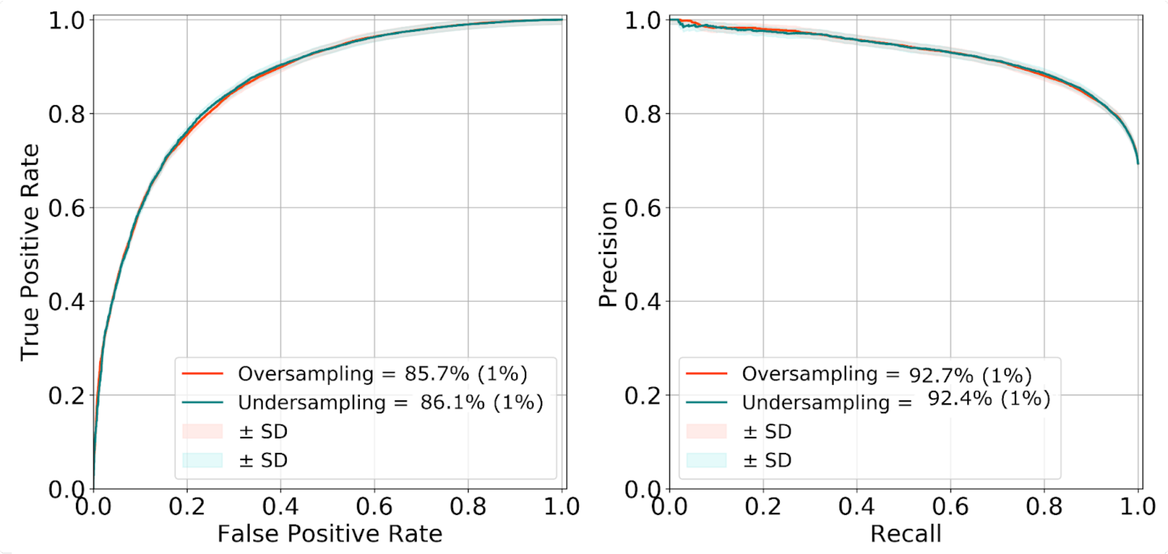


**Supplemental Material 9: 9a.) AUROC and 9b.) AUPRC for SIgnose with oversampling of the minority class and random undersampling of the majority class.**

**Supplemental Material 10:** SHAP decision plot for misclassified and correctly classified observations.

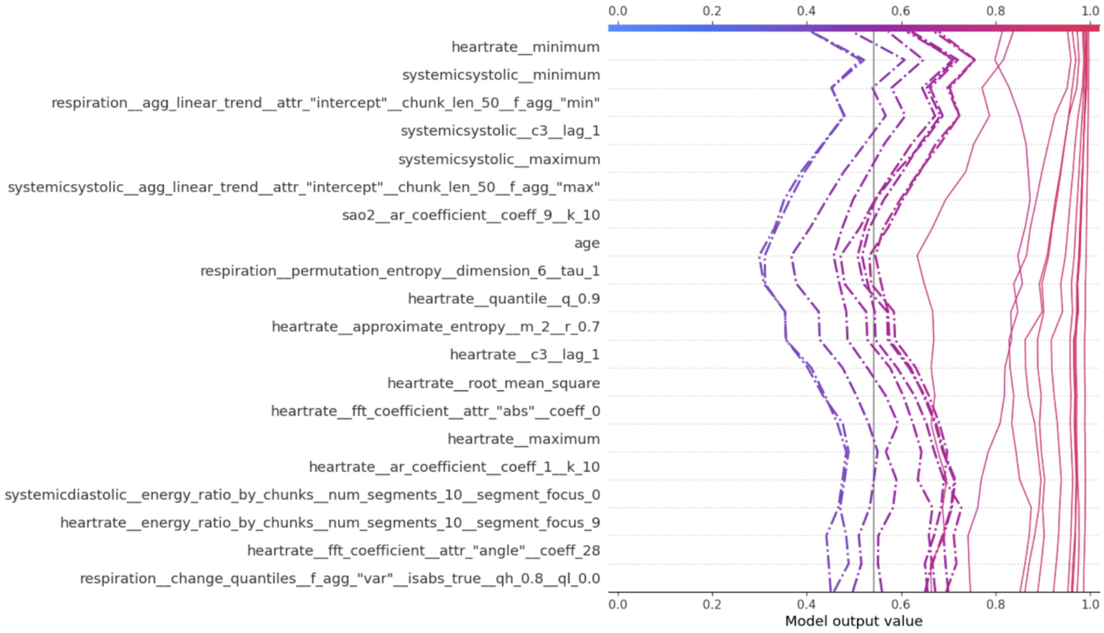


**Supplemental Material 10: SHAP decision plot for misclassified (highlighted as dashed lines) and correctly classified observations.**

**Supplemental Material 11:** Performance of *SIgnose* fine-tuned Model on different age groups, new settings' ICU types, and time since admission.

SIgnose was fine-tuned and tested on MIMIC-III data. Similarly, it was also fine-tuned and tested on SafeICU pediatric data. Models showed consistent recall and precision above 90% and 81%, respectively, across all age groups, except for the 30-40-year age group (Supplemental Material 11a). The model also consistently achieved recall above 90% across all ICU types, including the pediatric ICU (Supplemental Material 11b). In the MIMIC-III cohort, performance with time since admission reached recall above 90% from the ninth hour of admission (Supplemental Material 11c). In contrast, in SafeICU pediatric data, it remained consistently high, with a recall of 100% from the beginning (Supplemental Material 11d).

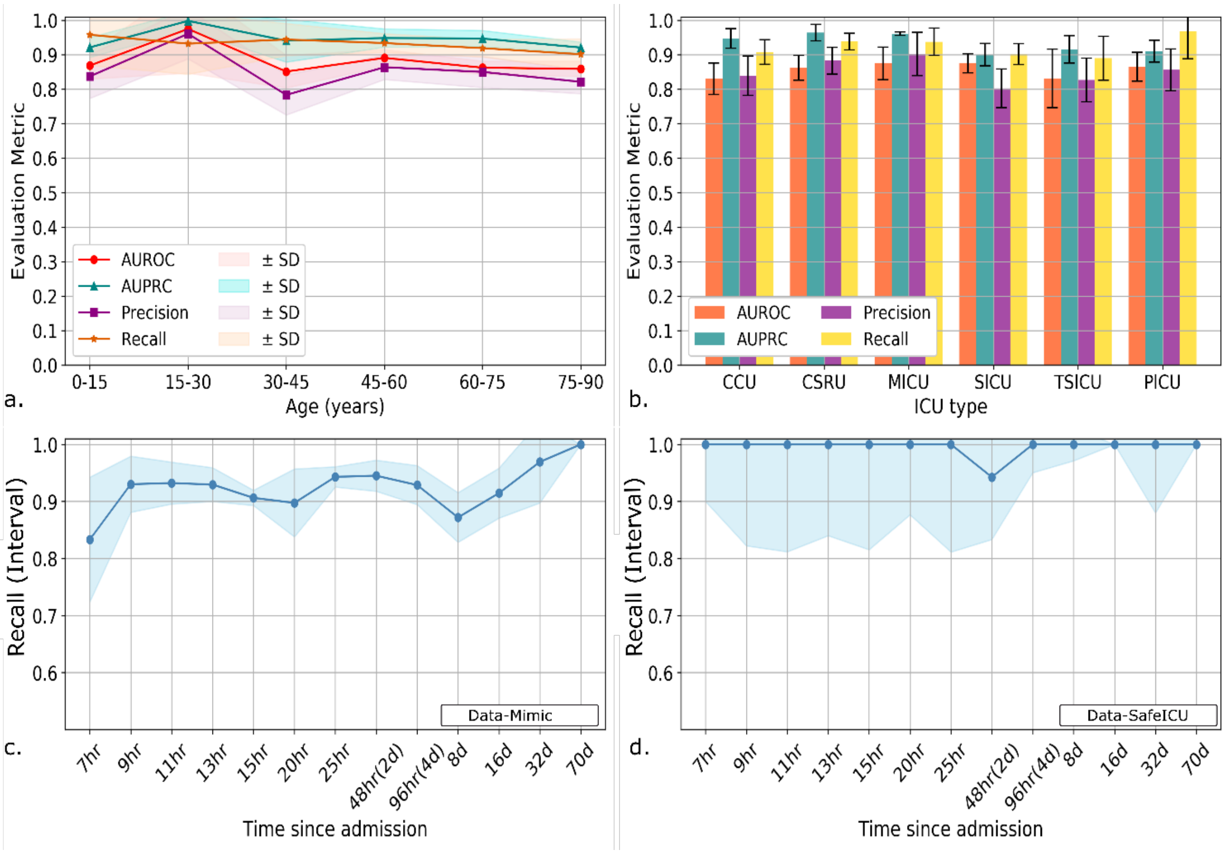


**Supplemental Material 11: 11a.) SIgnose performance parameters for external validation on different age groups (SafeICU pediatric age group is embedded along with MIMIC-III (adult age groups) data) 11b.) Performance of the fine-tuned model in different care units. 11c.) fine-tuned on MIMIC-III data, model recall for "time since admission" 11d.) fine-tuned on SafeICU pediatric data, model recall for "time since admission."**

\

**Supplemental Material 12:** Model performance across several diagnosis categories.

| Precision | Recall | AUC | AUPRC | Diagnosis | Sample (n patients) | Data |
|---|---|---|---|---|---|---|
| 0.9 | 0.82 | 0.82 | 0.88 | Hypertension | 993 | MIMIC |
| 0.94 | 0.85 | 0.83 | 0.92 | Heart failure | 507 | MIMIC |
| 0.92 | 0.84 | 0.81 | 0.9 | Atherosclerosis | 601 | MIMIC |
| 0.93 | 0.85 | 0.83 | 0.91 | Diabetes | 462 | MIMIC |
| 0.92 | 0.87 | 0.8 | 0.91 | Pneumonia | 448 | MIMIC |
| 0.91 | 0.82 | 0.84 | 0.89 | Malignant Neoplasm | 361 | MIMIC |
| 0.92 | 0.86 | 0.83 | 0.91 | Anemia | 442 | MIMIC |
| 0.91 | 0.84 | 0.8 | 0.9 | Coronary Artery | 553 | MIMIC |
| 0.93 | 0.87 | 0.83 | 0.92 | Atrial fibrillation | 467 | MIMIC |
| 0.92 | 0.87 | 0.81 | 0.91 | Acute kidney failure | 417 | MIMIC |
| 0.89 | 0.86 | 0.8 | 0.87 | Chronic kidney disease | 219 | MIMIC |
| 0.94 | 0.86 | 0.83 | 0.93 | Respiratory failure | 383 | MIMIC |
| 0.93 | 0.83 | 0.82 | 0.91 | Hemorrhage | 272 | MIMIC |
| 0.92 | 0.84 | 0.84 | 0.9 | Hypercholesterolemia | 291 | MIMIC |
| 0.93 | 0.83 | 0.82 | 0.91 | Urinary tract infection | 274 | MIMIC |
| 0.94 | 0.86 | 0.83 | 0.92 | Septal defect | 45 | SafeICU |
| 0.89 | 0.89 | 0.81 | 0.88 | Pneumonia | 38 | SafeICU |
| 0.96 | 0.92 | 0.79 | 0.95 | Acyanotic congenital heart disease | 27 | SafeICU |
| 0.89 | 0.85 | 0.59 | 0.89 | Shock | 23 | SafeICU |
| 0.94 | 0.88 | 0.84 | 0.93 | Hypertension | 22 | SafeICU |
| 0.91 | 0.83 | 0.85 | 0.88 | Congenital heart disease | 20 | SafeICU |
| 0.82 | 0.88 | 0.64 | 0.82 | Gastroenteritis | 21 | SafeICU |
| 1 | 0.93 | 1 | 1 | Sepsis | 14 | SafeICU |
| 1 | 0.9 | 0.95 | 0.99 | Lower respiratory tract infection | 12 | SafeICU |
| 1 | 1 | 1 | 1 | Congestive heart failure | 9 | SafeICU |

**Supplemental Material 13:**

To assess the potential of the large language model (LLM)-based embeddings, we employed Llama-3.1 (8B), an 8-billion-parameter decoder-only transformer model released by Meta. We used a 4-bit quantized model using the "unsloth" Python library. Structured 5-minute-interval vital signs data from the eICU database were converted into textual prompts encoding demographic and physiological information, which were then tokenized and passed through the Llama-3.1 backbone to extract fixed-length embeddings. These embeddings were subsequently used as input features for downstream classical machine learning classifiers. In selected experiments, parameter-efficient fine-tuning of the Llama-3.1-8B 4-bit model was performed using Low-Rank Adaptation (LoRA) on the source dataset before embedding extraction, while downstream classifiers were trained separately. All LLM experiments were conducted on 2 NVIDIA L40S GPUs.

**Supplemental Material 14:**

Application overview (SafeICU)

This application predicts the probability of abnormal shock index in critically ill patients within the next 8 hours. It is based on a machine-learning model prospectively validated in a pediatric ICU at AIIMS New Delhi.

Data requirements

The application requires 420 minutes of physiological time series data sampled at 5-minute resolution, including:

- Heart rate
- Arterial systolic blood pressure
- Arterial diastolic blood pressure
- Oxygen saturation
- Respiratory rate

Patient age and sex are additionally required as inputs.

Deployment

For reproducibility, the application is containerized using Docker. During review, access credentials, container identifiers, and source-code repositories are withheld to preserve anonymity. Source code is publicly available at https://github.com/tavlab-iiitd/SIgnose; Docker container images will be made available upon acceptance.

The container exposes a web-based interface that allows users to upload CSV-formatted physiological time series data and receive risk estimates.

Application workflow

1. Upload a CSV file containing the required physiological variables and demographic inputs.
2. The application automatically preprocesses the uploaded data, including variable identification and normalization.
3. The model outputs a probability score indicating the risk of an abnormal shock index over the prediction horizon, using a gradient-boosted decision-tree classifier trained on retrospective ICU data.

Intended use

This application is intended as a clinical decision-support tool for ICU settings and is not designed to replace clinical judgment. It should not be used outside critical care environments.
